# Cardiac Post-acute Sequelae symptoms of SARS-CoV-2 in Community-Dwelling Adults: Cross-sectional Study

**DOI:** 10.1101/2022.07.05.22277260

**Authors:** Oluwabunmi Ogungbe, Nisha A. Gilotra, Patricia M. Davidson, Jason E. Farley, Cheryl R. Dennison Himmelfarb, Wendy S. Post, Yvonne Commodore-Mensah

## Abstract

**Objective:** To examine risk factors for cardiac-related PASC in community-dwelling adults after acute coronavirus disease 2019 (COVID-19) infection.

**Methods:** We performed a cross-sectional analysis among adults who tested positive for COVID-19. Outcomes were self-reported cardiac-related PASC. We conducted stepwise multivariable logistic regression to assess association between the risk factors (existing cardiovascular disease, pre-existing conditions, days since positive test, COVID hospitalization, age, sex, education, income) and cardiac-related PASC.

**Results:** Among a sample of 442 adults, mean(±SD) age was 45.4 (16.2) years, 71% female, 13% Black; 46% reported pre-existing conditions; 23% had cardiovascular (CV) risk factors; and 4% had cardiovascular diseases (CVD). The prevalence of persistent cardiac-related symptoms and newly diagnosed cardiac conditions was 43% and 27%, respectively. The odds for cardiac-related PASC were 2.01 (95%CI: 1.27-3.17) higher in persons with underlying CV risk factors/CVD than in those without. The odds for cardiac-related PASC were higher among persons with underlying pre-existing conditions (adjusted odds ratio aOR: 2.00, 95% CI:1.28-3.10) and among those who were hospitalized (aOR: 3.03, 95%CI:1.58-5.83).

**Conclusions:** More than a third of persons with COVID-19 reported cardiac-related PASC symptoms. Risk factors for cardiac-related PASC symptoms include underlying CV risk factors/CVD, pre-existing conditions, increasing age, and COVID-19 hospitalization. COVID-19 may play an important role in worsening the prognosis of existing CV risk factors and increasing risk of complications.

**Key Messages:** *What is already known on this topic:* Infection with SARS-CoV-2 virus may lead to persistent cardiac symptoms indicative of cardiac injury. COVID-19 may play an important role in worsening the prognosis of existing CVD and pre-existing conditions.

*What this study adds:* Little is known about characterization of cardiac-related Post-Acute Sequelae of SARS-CoV-2 Infection (PASC) and recovery in community-dwelling adults. Hence, this study provides further evidence on the burden of possible cardiac-related PASC symptoms and diagnosed cardiac conditions. The findings also suggest that underlying CVD, pre-existing conditions, older age, and COVID-19 hospitalization, may be risk factors for persistent cardiac-related PASC symptoms.

*How this study might affect research, practice or policy:* These results underscore urgent needs for coordinated efforts directed at resource allocation and optimization of primary care for persons with cardiac PASC and for prevention of resulting CV events and complications

## Introduction

As of April 2022, the cumulative number of Coronavirus Disease 2019 (COVID-19) cases was 493 million worldwide and 80 million in the US, resulting in over 6 million deaths globally and almost a million deaths in the US.^1^ Post-acute sequelae of SARS-CoV-2 infection (PASC), aka post-acute COVID syndrome or Long-COVID, is an emerging and sometimes debilitating health condition in persons with prior COVID-19.^2^ The National Institutes of Health defines PASC as the persistence of symptoms or development of sequelae beyond three weeks from the onset of acute symptoms of SARS-CoV-2 infection.^3^

Beyond 30 days of acute illness, patients who have recovered from COVID-19 exhibit a wide range of cardiopulmonary and neurological clinical manifestations, increased healthcare utilization, generally report poor wellbeing and sometimes experience late mortality.^4^ Reports have documented that the most frequent cardiopulmonary PASC symptoms include: dyspnea (52%), post-exertional fatigue (42%), chest pain/pressure (48%), and palpitations (44%).^5^ Tachycardia (58%), orthostatic intolerance and syncope (41%), signs of postural orthostatic tachycardiac syndrome (POTS) (69%), inappropriate sinus tachycardia (20%), and other signs of cardiovascular autonomic dysfunction are also common in PASC.^2^ These symptoms were observed up to a median of 6 months after symptoms onset among recovered hospitalized patients.^6^ The more frequent diagnosis includes the following: myocardial injury (myocardial fibrosis or myocarditis) in 9-78%^7^ including lower left ventricular ejection fraction and higher left ventricular volumes,^8^ myocardial involvement (15%) by cardiac MRI, nonischemic cardiomyopathy (5%), new ischemia (3%), new supraventricular tachycardia (2%), coronary vasospasm (2%), and new atrial fibrillation (2%).^5^ Cardiopulmonary exercise testing among patients reporting PASC found excessive ventilator response (88%), circulatory impairment (58%), and a higher prevalence of symptoms consistent with chronic fatigue syndrome (46%).^9^

While the prevalence of these persistent symptoms continues to be documented among hospitalized persons, far fewer studies have been conducted among patients who were never hospitalized or categorized as having “mild” symptoms. Among non-hospitalized patients, a study reported that 68.7% experienced at least one symptom 30 days after acute infection, and 73% reported symptoms ≥60 days of follow-up.^10^ Various diagnoses, including new-onset diabetes mellitus, neurological and cardiac disorders, are also emerging among patients who were never hospitalized for COVID-19.^11^ PASC can lead to long-term cardiovascular morbidity and disability among persons who continue to experience these symptoms.^12^ Although several health systems now have functioning post-COVID clinics, several gaps remain and a dearth of explanation for these symptoms, and inadequacy of clinical management modalities, including public health strategies, to address PASC syndrome. In this study, we present results from a study of a majority of community-dwelling cohorts with previous COVID-19 infection. We examine persistent cardiac-related PASC symptoms in patients after acute COVID-19 illness.

## Methods

The Cardiac PASC study is an ongoing prospective study that began in 2021. This nested sub-study includes a cross-sectional evaluation of baseline data from adults enrolled in the Johns Hopkins COVID-19 HOPE registry,^13^ which is a centralized database that contains information on participants interested in COVID-19 studies at Johns Hopkins University. HOPE registry participants are predominantly from the Baltimore-Washington DC metropolitan area who had COVID-19 testing at any of the Johns Hopkins health system sites.

### Study Sample

Participants from the HOPE registry were eligible for the current study if they were (1) at least 18 years old; (2) had ever tested positive for COVID-19, ascertained through electronic medical record (EMR) test records of positive SARS-CoV-2 reverse transcriptase-polymerase chain reaction (RT-PCR) tests; (3) currently enrolled in the HOPE registry, (4) provided informed consent, and (5) willing to participate in the study. Exclusion criteria were: (1) adult participants who are unable to provide informed consent; (2) pregnant persons (ascertained through self-report). We invited participants to join the study in weekly batches through automated invitations generated in REDCap.

#### Sampling

We used a stratified sampling procedure in this study. The HOPE registry has an overrepresentation of non-Hispanic White adults (80%); hence, in this study, we applied stratified sampling by race and ethnicity to allow oversampling for persons who do not identify as non-Hispanic White. We employed this sampling strategy to obtain a sample reflective of the population-level race/ethnicity distribution in the United States. After applying the pre-specified eligibility criteria to participants in the HOPE registry, we obtained the sample frame stratified by race/ethnicity with a total of 7 strata (Race: White, Black/African American, Asian, American Indian/Alaska Native, other races; Ethnicity: Hispanic, Non-Hispanic). We used disproportionate sampling within the strata for persons who identified as Black/African American, Asian, American Indian/Alaska Native, other races, and Hispanic, inviting 100% of them to join the study due to an initial lower response rate from persons within these strata. We applied proportionate simple random sampling for the non-Hispanic White Stratum, inviting 60% of them to join the study (Figure 1) since the initial response rate was higher among this population.

**Figure 1:**
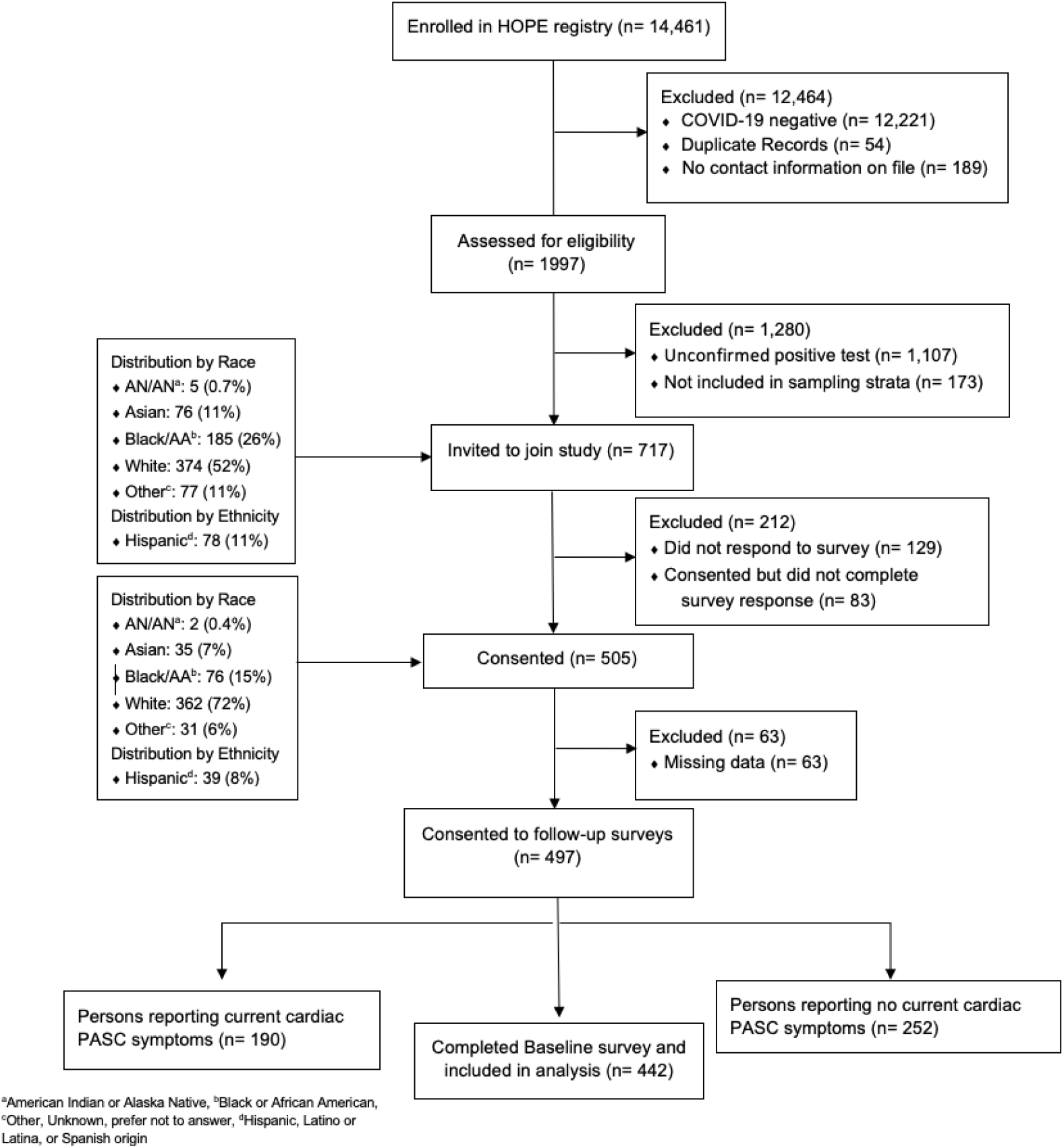
Flow diagram demonstrating inclusion and exclusion of participants in the study

#### Recruitment

We sent email invitations generated directly from REDCap and text invitations using the REDCap-Twillio connection.^14^ The invitations contained information about the study, describing the study as “*lingering heart-related symptoms from COVID-19*,” and survey entry links unique to each participant, which could not be shared. This invitation also included information about the follow-up surveys. We enrolled participants between November 2021 and January 2022.

The survey response rate was 70%; thus, we included a sample of 449 participants in this analysis. At survey completion, participants received a $10 digital gift card in compensation through an automated process in REDCap. Figure 1 shows a flow diagram of the participants included in this analysis.

### Data Collection and measurement

Baseline information on COVID-19 was obtained by self-reported surveys. The baseline surveys included questions on participants’ sociodemographic characteristics, COVID-19 testing, exposure, and vaccinations, pre-existing conditions, persistent cardiac-related symptoms, cardiac complications, fatigue, and overall health. The baseline survey took approximately 10-15 minutes to complete. In addition, we ascertained COVID-19 testing date through EMR test records in the HOPE registry. We assessed baseline cardiovascular disease or risk factors through self-report.

#### Pre-existing CV risk factors, CVD, and other conditions

We assessed pre-existing CVD by asking participants: “*Before the COVID-19 pandemic (or before March 12, 2020), have you been diagnosed with any of the following? 1. hypertension, 2. High cholesterol, 3. coronary artery disease, 4. chronic ischemic disease, 5. heart failure, 6. valvular heart disease, 7. atrial fibrillation; etc*.*”* Participants were encouraged to list other diagnosed CVD previously not listed. For other pre-existing conditions, we listed chronic diseases such as stroke, diabetes mellitus, kidney disease, cancer, etc., and asked participants to list other conditions they may have had prior to COVID-19.

#### PASC Symptoms and newly diagnosed cardiac conditions

At the time of survey administration, there were few validated tools for assessing PASC symptoms. Thus, we developed questionnaires on PASC symptoms based on symptoms reported in systematic reviews.^15, 16^ These symptoms were categorized based on the body system affected: general symptoms (fatigue or tiredness, night sweats, etc.), respiratory symptoms (shortness of breath, cold feelings in lungs, etc.), cardiovascular (persistent chest pain, tachycardia, etc.), musculoskeletal (joint pain, muscle aches, etc.), neurological (difficulty concentrating, tremors, etc.), gastrointestinal (heartburn, diarrhea), psychological (anxiety, sadness, etc.), eyes (blurry vision, dry eyes, etc.), ear, nose and throat (tinnitus, sore throat, etc.), skin and hair (rash, hair loss, etc.). Participants also listed additional symptoms not previously listed in either of these categories. We asked participants to indicate whether they had received any new cardiac diagnoses: *“Since testing positive for COVID-19, has your healthcare provider newly diagnosed you with any of the following? 1. hypertension, 2. tachycardia, 3. myocarditis, 4. cardiomyopathy. 5. myocardial injury, 6. atrial fibrillation, 7. pericarditis, 8. myocardial infarction, 9. heart block, 10. heart failure, 11. pulmonary embolism, 12. deep vein thrombosis, 13. stroke, 14. acute kidney disease, 15. postural orthostatic tachycardia syndrome – POTS”*. Participants had the option to input new diagnoses that were not listed. We defined each of these diagnoses in participant-friendly language at appropriate health literacy levels.

#### Fatigue Short Form (FACIT-Fatigue)

To assess self-reported levels of fatigue, we used the Functional Assessment of Chronic Illness Therapy (FACIT) Fatigue Scale (Version 4), which is one of many scales in the FACIT Measurement System, a collection of health-related quality of life questionnaires targeted toward managing chronic illness. The FACIT Fatigue Scale is a short, 13-item tool that measures individuals’ levels of fatigue during their usual daily activities over the past week. We measured fatigue levels on a four-point Likert scale (4 = not at all fatigued to 0 = very much fatigued); total scores ranged from 0-52; lower scores indicated severe fatigue.^17^ Items 7 and 8 on energy and ability to do usual activities were reverse-coded, and we also dichotomized the sum score using a recommended cut point of 34.^18^

#### Global health (PROMIS Scale)

We administered the PROMIS Scale Global Health (PROMIS-GH) v1.2, a 10-item scale representing the following five core health domains: physical health, pain, fatigue, mental health, social health, and overall health.^19^ Each item was scored on a 5-point Likert scale (except for item 7) that was scored on an 11-point numerical scale and recoded to a 5-points scale (1 = None to 5 = Very severe). Items 8 and 10 were reverse-coded. For the analysis, we obtained two sum scores. The General Mental Health (GMH) score for mental health was calculated from items 2, 4, 5, and 10; the General Physical Health (GPH) score for physical health was calculated from items 3, 6, 7, and 8. Items 1 on general health, item 9 on ability to carry out social activities were analyzed as single items and not included in the sum scored. Higher scores indicated better global mental/physical health.

### Outcomes

The outcome of the study was self-report of possible cardiac-related PASC beyond two weeks of initial acute COVID-19 infection and at the time of survey administration. We asked participants to indicate which COVID-related symptoms they experienced within ≥3 weeks of acute COVID-19 infection, whether they were experiencing any of these symptoms at enrollment, and the duration of their symptoms. Cardiac-related PASC, the primary outcome of this study, was defined as the presence of at least one cardiac PASC symptom ≥3 weeks after a positive COVID-19 test and at survey administration. Symptoms included sharp or sudden chest pain, tachycardia or higher than normal pulse rate, feeling faint, or heart palpitations.^20^ Due to the non-specificity of fatigue symptoms, we excluded fatigue from our definition of cardiac-related PASC. We calculated time since infection as the interval between the date of the COIVD-19 test and the survey consent date. We also assessed new diagnoses of cardiovascular diseases through self-report based on participants’ answers to the question: “*Since testing positive for COVID-19, has your healthcare provider newly diagnosed you with any of the following?”* We computed the FACIT fatigue outcome as a sum score of the FACIT fatigue scale; the physical, mental, and social functioning was computed from the Global Health PROMIS scale.

### Patient and public involvement

Patients or the public were not involved in the design, or conduct, or reporting, or dissemination plans of our research.

### Ethical Statement

Ethical approval for the present study was obtained from the Johns Hopkins Medicine Institutional Review Board (IRB00299548). Informed consent was obtained from each participant.

### Statistical Analysis

We performed a cross-sectional analysis of baseline data from an ongoing cohort study of persons who had tested positive for COVID-19. Following a missing data analysis, and observations for the outcome variable determined to be missing at random (MAR) were excluded from the final analysis (Figure 1 in the Supplement). Sample characteristics were visually summarized and assessed to identify distributions, outliers, and missing data patterns. We reported categorical variables in frequencies and percentages and continuous variables in mean and standard deviation or median (interquartile range or range) depending on the normalcy of distribution. Sociodemographic characteristics were summarized and stratified by cardiac-related PASC symptoms at enrollment outcome. The prevalence of the most reported cardiac-related PASC symptoms were summarized. Using test of proportions, we compared the prevalence of these symptoms at the 2-weeks post-acute COVID-19 and on enrollment. A bivariate analysis was performed to examine what predictor variables were most associated with current reports of PASC and cardiac-related PASC symptoms.

Report of cardiac-related PASC symptoms ≥ 3 weeks after positive COVID-19 test, current report of PASC symptoms, report of underlying cardiovascular symptoms, and report of pre-existing conditions were examined as dichotomous variables (yes/no). We examined the association between underlying CV risk factors/CVD, and other pre-existing conditions and reports of cardiac-related PASC symptoms in crude logistic regression models. We then performed adjusted logistic regression analyses, adjusting for age, gender, race, education, household income, and time since COVID-19 infection. We then performed stepwise multivariable logistic regression with backward selection to assess the association between underlying cardiovascular disease or risk factors and report of current cardiac-related PASC symptoms to permit statistical selection of the best predictor variables. We considered predictors with *p*<0.2 for inclusion in the stepwise multivariable logistic regression performed for the outcomes of this study.

We assessed the association between fatigue and cardiac-related PASC symptoms. We derived a sum score based on the FACIT fatigue scale and, using summary statistics techniques, we summarized the total sum score stratified by current cardiac-related PASC symptoms. We then dichotomized the sum fatigue score into two categories (severe fatigue/less severe fatigue), using the recommended 34-point cut point.^18^ We performed multivariable logistic regression analyses for the relationship between fatigue and cardiac-related PASC symptoms. We obtained sum scores for mental and physical health functioning separately and assessed for differences in means scores using *T*-tests by report of cardiac-related PASC symptoms.

To further examine the temporal presentation of cardiac-related PASC symptoms, time since COVID-19 infection was divided into categories of 6 months (i.e., 0-5 months, 6-11 months, 12-17 months, and ≥18 months), and we plotted the prevalence of current cardiac-related PASC symptoms for each of these time bins. We also reported the prevalence of newly diagnosed conditions after recovery from COVID-19, stratified by time since infection (dichotomized into <18 months and ≥18 months). *P*<0.05 was considered statistically significant. Data analyses were conducted in Stata I/C version 16.1, and R version 2021.09.0 (R project or statistical computing).

## Results

As of November 2021, 1,997 adults with previous COVID-19 infection from the HOPE registry were eligible for this study and were sent 717 survey invitations. A total of 505 persons (70%) joined the study; after accounting for incomplete outcomes responses and missing data, we included a sample of 442 (88%) in the analyses (**Figure 1**). Sample sociodemographic characteristics stratified by cardiac-related PASC symptoms are reported in **Table 1**. At enrollment, the mean (SD) age was 45.4 (**±**16.2) years, 71% were female, 13% were Black, 29% had underlying CV risk factors (23%) and CVD (4%) prior to COVID-19 infection. In the sample, 43% reported current cardiac-related PASC symptoms; only 12% reported being hospitalized for COVID-19, and Median (IQR) time since infection was 12.4 (10.0-15.2) months. About 10.5% were unvaccinated, and among those who had received at least one dose of the COVID-19 vaccine (400, 90.5%), 56.7% had received a booster dose. About 73% had received the first vaccine dose before SARS-CoV-2 positive test.

**Table 1.** Sociodemographic characteristics stratified by report of current cardiac-related PASC symptoms (N = 442)

| Characteristics, M(±SD) / n(%) | Total<br>(N = 442) | Cardiac PASC symptoms | | P-value |
| --- | --- | --- | --- | --- |
|  |  | No<br>(N= 252) | Yes<br>(N= 190) |  |
| **Mean(SD)/n(%)** |  |  |  |  |
| **Age, years** | 45.4 (16.2) | 43.2 (16.0) | 48.2 (16.1) | **0.002** |
| **Female<sup>a</sup>** | 311 (70.8) | 171 (68.1) | 140 (73.7) | 0.116 |
| **Underlying CV risk factors and CVD<sup>b</sup>** | 129 (29.2) | 56 (22.2) | 73 (38.4) | **<0.001** |
| **Pre-existing conditions<sup>c</sup>** | 203 (45.9) | 95 (37.7) | 108 (56.8) | **<0.001** |
| **Months since COVID-19 infection<sup>d</sup>** | 12.0 (5.3) | 11.3 (5.3) | 12.8 (5.3) | **0.005** |
| **Race/Ethnicity** |  |  |  | 0.255 |
| White | 322 (72.9) | 190 (75.4) | 132 (69.5) |  |
| Black/African American | 55 (12.6) | 27 (10.7) | 28 (14.7) |  |
| Asian | 33 (7.5) | 21 (8.3) | 12 (6.3) |  |
| Other | 16 (3.6) | 8 (3.2) | 8 (4.2) |  |
| Hispanic ethnicity<sup>e</sup> | 31 (7.1) | 12 (4.8) | 19(10.0) | 0.154 |
| **Educational status** |  |  |  | 0.331 |
| High School diploma/≤GED | 27 (6.1) | 11 (4.4) | 16 (8.4) | |
| Some college | 100 (22.7) | 57 (22.7) | 43 (22.6) |  |
| Bachelor's degree | 152 (34.5) | 91 (36.3) | 61 (32.1) |  |
| Graduate degree | 162 (36.7) | 92 (36.7) | 70 (36.8) |  |
| **Household Income** |  |  |  | 0.535 |
| ≤\$39,999 | 64 (14.7) | 31 (12.5) | 33 (17.5) | |
| \$40,000 – \$69,999 | 68 (15.6) | 37 (14.9) | 31 (16.4) | |
| \$70,000 – \$99,999 | 77 (17.6) | 43 (17.3) | 34 (18.0) | |
| ≥\$100,000 | 183 (41.9) | 111 (44.8) | 72 (38.1) | |
| **Employed<sup>f</sup>** | 307 (69.6) | 190 (76.3) | 117 (62.6) | 0.002 |
| **COVID-19 hospitalization** |  |  |  |  |
| Hospitalized, n (%) | 51 (11.5) | 15 (6.0) | 36 (19.0) | **0.001** |
| Days on admission, Median (Range) | 5 (1-117) | 4 (1-42) | 6 (1-117) | 0.492 |
| **COVID-19 Vaccination, n (%)** |  |  |  |  |
| At least one dose | 400 (90.5) | 232 (92.1) | 168 (88.4) | 0.164 |
| Two doses | 372 (84.2) | 218 (86.5) | 154 (81.1) | 0.120 |
| Booster dose | 230 (57.6) | 138 (59.5) | 92 (55.1) | 0.453 |
| Vaccine prior to SARS-CoV-2 infection<sup>g</sup> | 324 (73.3) | 179 (71.0) | 145 (76.3) | 0.214 |
| **Healthcare Worker<sup>h</sup>** | 146 (33.3) | 81 (32.7) | 65 (34.2) | 0.733 |

Ref: <sup>a</sup>Male; <sup>b</sup>CV risk factors included hypertension and dyslipidemia; CVD included coronary artery disease, heart failure, atrial fibrillation, chronic ischemic heart diseases, myocardial infarction, valvular heart disease, etc.; <sup>c</sup>Pre-existing conditions prior to COVID-19 infection (cardiovascular diseases, asthma, cancer, kidney disease, irritable bowel syndrome etc.); <sup>d</sup>Median (IQR) months since infection was 12.4 (10.0-15.2) months; <sup>e</sup>Non-Hispanic; <sup>f</sup>Not employed; <sup>g</sup>Received First dose of COVID-19 vaccine before report of positive SARS-CoV-2 infection; <sup>h</sup>Non-healthcare worker.
M(±SD) / n(%): Mean (±Standard Deviation)/frequency (percent). BMI: Body mass index. GED: General Education Development qualification Bold: Statistically significant at <0.05. *P*-values were estimated using t-tests for differences in means and chi-square test statistics for differences in proportions.

### Prevalence of cardiac-related PASC symptoms

Almost 52% of the sample (228/442) reported possible cardiac-related PASC symptoms lasting 3 weeks or more after acute infection; 43% reported symptoms (190/442) on enrollment. We examined the prevalence of each specific symptom comparing the 2-weeks post-acute COVID and current experience of these symptoms (**Figure 2**): heart palpitations was the most reported symptom (22.6% vs. 14.5%, *p*<0.05), then tachycardia (20.6% vs. 12.9%, *p*<0.05).

**Figure 2:**
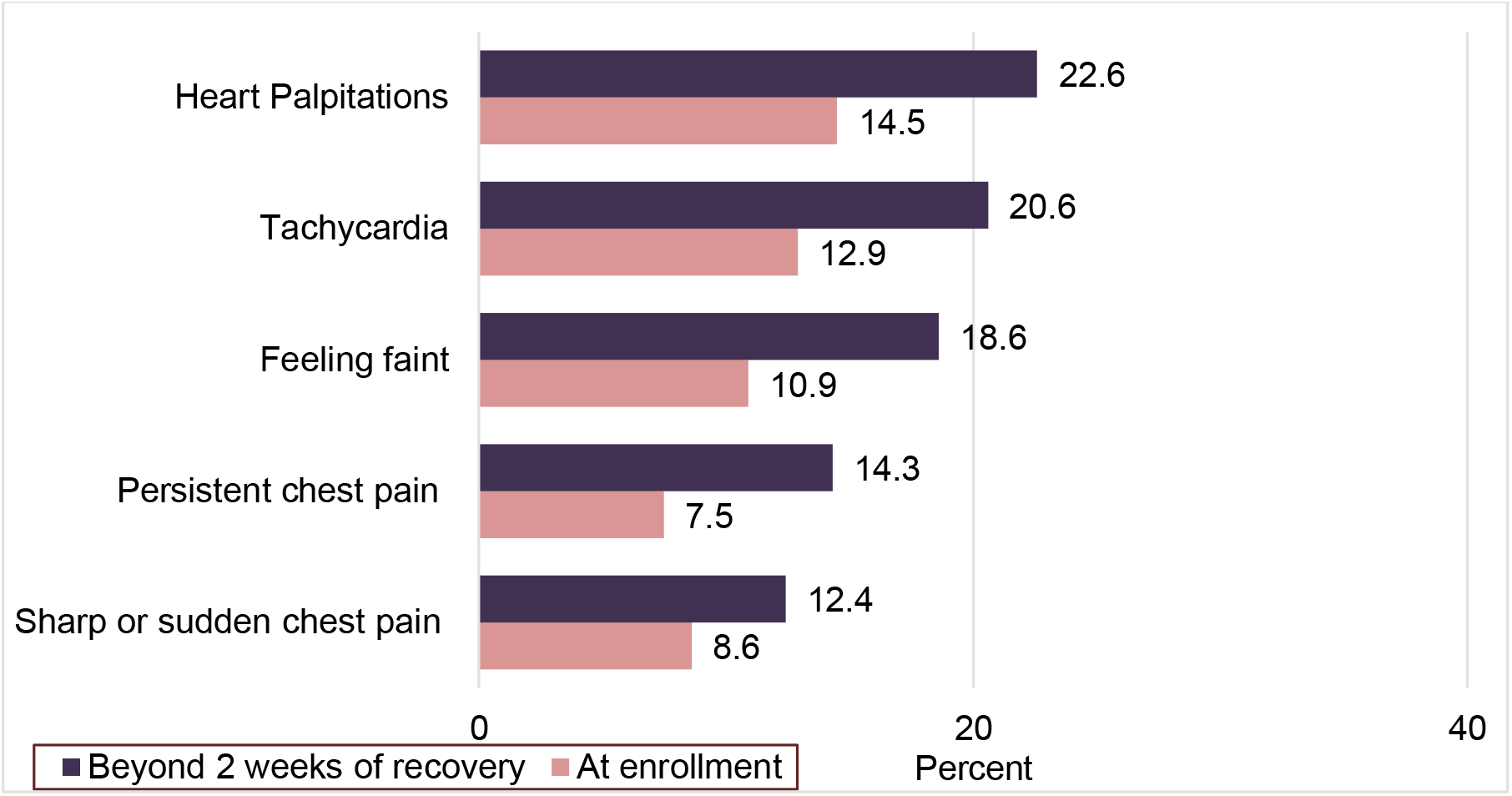
Prevalence of cardiac-related PASC symptoms, comparing beyond 2 weeks post-acute COVID and at survey administration (N = 442)

Feeling faint was also prevalent (18.6% vs. 10.9%, *p*<0.05), followed persistent chest pain (14.3% vs. 7.5%, *p*<0.05), and sharp chest pain (12.4% vs. 8.6%, *p*=0.062). The majority of these symptoms improved between the time of immediate post-acute infection (≥3 weeks) and at survey administration (mean of 12 months). Other general symptoms most reported were fatigue (42.3%), joint pain (26.7%), muscle pain (24.2%), shortness of breath on exertion (20.4%), and activity intolerance (17.7%).

### Risk factors for cardiac-related PASC symptoms

The univariable analyses showed statistically significant association between underlying CVD, pre-existing conditions, age, COVID-19 hospitalizations, and cardiac-related PASC symptoms, comparing less than 18 months to greater than 18 months since COVID-19 infection (**Figure 3**). Prevalence of cardiac-related PASC by time since COVID-19 infection categories (0-5 months; 6-11 months; 12-17 months and ≥18 months) showed a sustained high prevalence of these symptoms after 12 months of recovery (**Supplemental Figure 1**).

**Figure 3:**
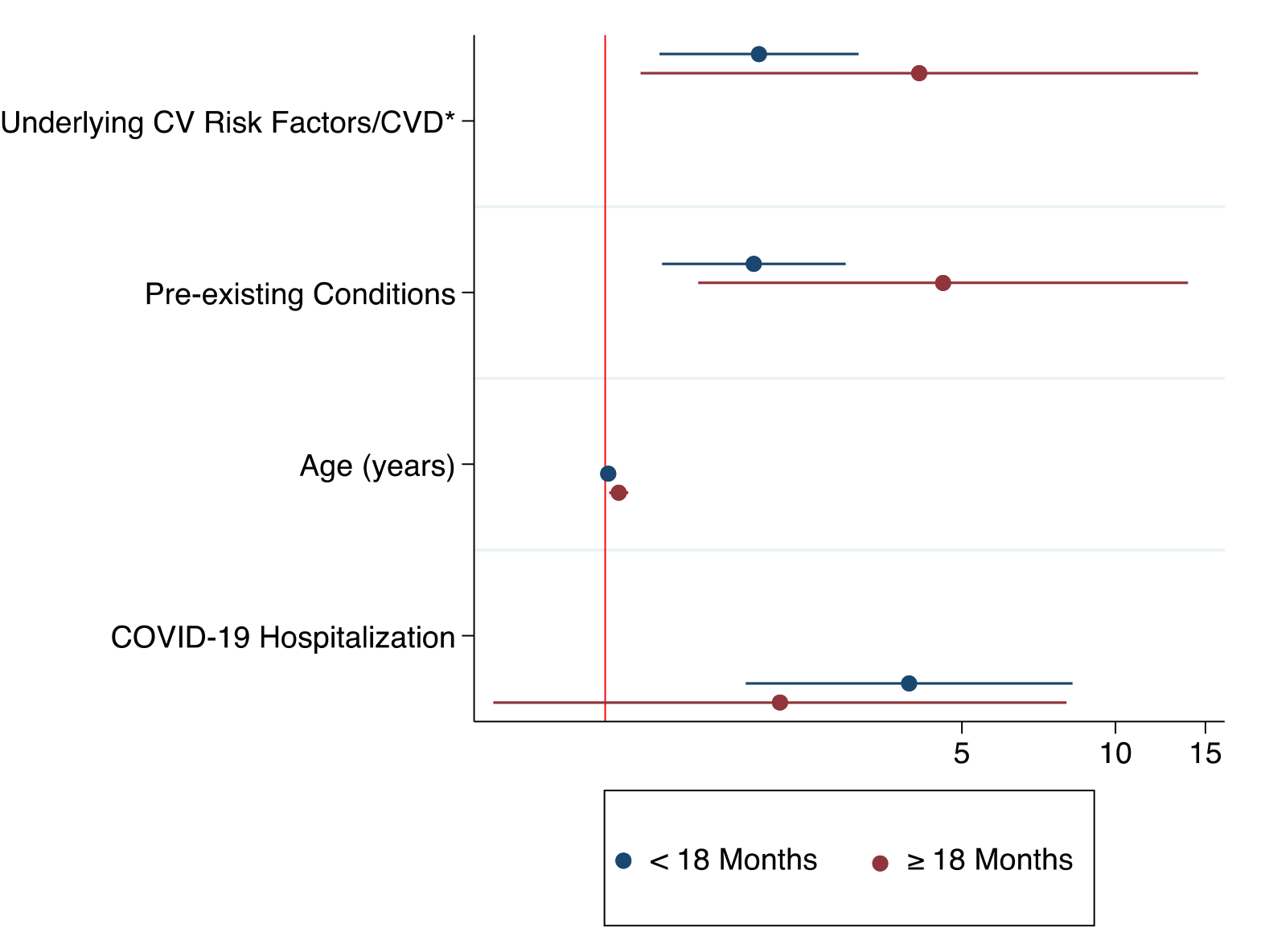
Univariate analysis for factors associated with current cardiac-related PASC symptoms, stratified by months since positive test (<18 months vs. ≥18 months) ^*^CV risk factors included hypertension and dyslipidemia; CVD included coronary artery disease, heart failure, atrial fibrillation, chronic ischemic heart diseases, myocardial infarction, valvular heart disease, etc. CV = Cardiovascular, CVD = cardiovascular disease

**Table 2** presents results from the univariate and multivariable logistic regression analyses. After adjusting for age, race, income, education, and days since a positive test, the odds of cardiac-related PASC symptoms were 2 times higher in persons with underlying cardiovascular disease (aOR: 2.01 95%CI:1.27-3.17), and in persons with existing pre-existing conditions (aOR: 2.00, 95% CI:1.28-3.10). Every one-year increase in age was associated with a 2% increase in odds of reporting cardiac-related PASC (aOR: 1.02 95%CI:1.01-1.03); ≥18 months since COVID-19 test (aOR: 2.01 95%CI:1.15-3.61) also showed 2 times higher odds. Persons hospitalized for COVID-19 were more likely than those who were not hospitalized for COVID-19 to report cardiac-related PASC symptoms (aOR: 3.03 95%CI:1.58-5.83).

**Table 2:** Association between risk factors and self-report of cardiac-related PASC symptoms

| Risk factors | Cardiac-related PASC at enrollment |  |
| --- | --- | --- |
|  | OR (95%CI) | aOR (95% CI) |
| **Pre-existing CV risk factors/CVD<sup>a</sup>** |  |  |
| No underlying CV risk factors/CVD (*ref.*) | 1.00 | 1.00 |
| Presence of underlying CV risk factors/CVD | 2.18 (1.44-3.31) | 2.01 (1.27-3.17) |
| **Pre-existing conditions<sup>b</sup>** |  |  |
| No pre-existing conditions (*ref.*) | 1.00 | 1.00 |
| Pre-existing Conditions | 2.17 (1.48-3.20) | 2.00 (1.28-3.10) |
| **Age (per year)** | 1.02 (1.01-1.03) | 1.02 (1.01-1.03) |
| **COVID-19 hospitalization** |  |  |
| Not Hospitalized for COVID-19 (*ref.*) | 1.00 | 1.00 |
| Hospitalized for COVID-19 | 3.69 (1.96-6.97) | 3.03 (1.58-5.83) |

CI = confidence interval, kg/m<sup>2</sup> = kilogram, aOR = adjusted odds ratio, *ref.* = reference group, CV = Cardiovascular, CVD = cardiovascular disease
<sup>a</sup> CV risk factors included hypertension and dyslipidemia; CVD included coronary artery disease, heart failure, atrial fibrillation, chronic ischemic heart diseases, myocardial infarction, valvular heart disease, etc.
<sup>b</sup>Pre-existing conditions prior to COVID-19 infection (cardiovascular diseases, asthma, cancer, kidney disease, irritable bowel syndrome, etc.).
Crude and adjusted odds ratios and 95% confidence intervals were estimated using multivariable logistic regression models.
Stepwise, final model: age, income, race, education, days since positive test.

### Newly diagnosed cardiac conditions and risk factors

Over a quarter of the participants (26.9%, 119/442) reported a newly diagnosed cardiac condition after recovery from acute COVID-19. Of these participants, 20% reported newly diagnosed hypertension, 24% reported tachycardia, and 13% reported new diagnoses of postural orthostatic tachycardia syndrome (POTS) by a healthcare provider (**Supplemental Table 1**). Persons who had a ≥18 months recovery from acute COVID-19 infection were more likely to report diagnosis of tachycardia (*p*<0.05) and myocarditis (*p*<0.05).

### Fatigue and global health functioning

Fatigue was the most reported general symptom (42.3%). The overall mean fatigue score was 26.4(± 13.0); persons reporting possible cardiac-related PASC were more likely than those who no longer had symptoms to report fatigue (Mean difference: -13.11 95%CI: -15.25, -10.98, *p*<0.001). Using the recommended 34-point cut-off point,^18^ 88% of the participants reported experiencing fatigue. After adjusting for age, sex, race, education, and income, the odds of reporting fatigue were higher (aOR: 9.32, 95%CI: 5.54-15.66) among those with current cardiac-related PASC symptoms compared to those without. This higher odd was also similar for hospitalized patients (aOR: 4.06, 95%CI: 2.12-7.78). We assessed global functioning using the PROMIS Global Health Scale; the mean score for mental health functioning was lower among those reporting cardiac-related PASC symptoms at enrollment than among those who did not (Mean difference: -1.90 95%CI: -2.27, -1.53, *p*<0.001). Mean physical health functioning was also lower in the group reporting cardiac-related PASC symptoms (Mean difference: -1.44 95%CI: -1.87, -1.01, *p*<0.001).

## Discussion

The spectrum of cardiac-related PASC—particularly in community-dwelling persons with acute COVID-19 infection not requiring hospitalization—and its CV implications remain unclear. In this study, more than a third of persons with COVID-19 continued to report possible cardiac-related PASC symptoms over a mean duration of 12 months, emphasizing the longer-term impact of the virus, and more than a quarter reported diagnosis of new-onset cardiac risk factors and conditions post-acute COVID-19 infection. The most frequently reported cardiac-related PASC symptoms were heart palpitations, tachycardia, feeling faint, and chest pain. Risk factors associated with reporting of cardiac-related PASC symptoms were older age, underlying pre-existing conditions including CV risk factors and CVD, and hospitalization for acute COVID-19 infection. We also found that persons reporting possible cardiac-related PASC symptoms were more likely to report fatigue, worse general physical and mental functioning, and lower quality of life.

We found cardiac-related PASC to be prevalent with significant symptoms lasting for months post infection. Cardiopulmonary symptoms have been reported in up to 89% of non-hospitalized patients following acute COVID infection.^21^ Our findings provide further evidence of persisting symptoms in non-hospitalized patients: 52% reported possible cardiac-related PASC symptoms ≥3 weeks of COVID-19 positive test; 43% reported ongoing cardiac-related PASC symptoms at the time of study survey administration, despite a mean of 12 months post-acute infection. This is comparable with reports from other studies which have also reported persistent symptoms, including increased risk for incident cardiovascular disease greater than 12 months after acute infection.^2,22,23^ According to a recent American College of Cardiology (ACC) report, it remains unclear whether there are major distinctions in the underlying mechanisms, evaluation or management comparing cardiac-related PASC symptoms in the immediate post-acute period and symptoms extending beyond 12 months.^24^

Studies have reported structural injury to the heart months following acute recovery from COVID-19 in non-hospitalized patients considered low risk for PASC or COVID complications.^25^ Subsequently, evidence is emerging on the substantial burden of PASC among community-dwelling persons who represent the majority of persons with COVID-19.^26^ Our results are consistent with reports on enduring PASC in non-hospitalized patients,^27^ including reports of fatigue as the most common PASC.^28^ In our analyses, we observed that among persons with cardiac-related PASC symptoms, those who were hospitalized, 88% reported severe fatigue, lasting well over 6 months since recovery from acute COVID-19 infection. This poses significant implications on individuals, including inability to work or perform daily activities, job loss, severe financial slides, and strain on the work force at the population level. As such, there are needs for improved access to care and increased awareness of cardiac-related PASC symptoms in persons who have had COVID-19. As such, there are needs for improved access to care and increased awareness of cardiac-related PASC symptoms in persons who have had COVID-19.

We also found increased prevalence of self-reported diagnoses of CV risk factors and CVD. Over a quarter of participants in our study reported diagnoses of new CV risk factors and cardiac conditions such as hypertension after COVID-19 infection; however, this finding should be interpreted with caution due to the variability in the range of symptoms and clinical presentation of cardiac-related PASC and newly diagnosed conditions. Among 73,455 non-hospitalized patients in the US Department of Veterans Affairs health services, there was a high risk of incident CVD beyond 30 days of infection (hypertension, HR 15.18 [95%CI 11.53–18.62]; cardiac dysrhythmias, HR 8.41 [95%CI 7.18–9.53]), circulatory signs and symptoms, HR 6.65 [95%CI 5.18–8.01]; chest pain, HR 10.08 [95%CI 8.63–11.42]; coronary atherosclerosis, HR 4.38 [95%CI 2.96–5.67]; and heart failure, HR 3.94 [95%CI 2.97–4.80]).^4^ The most reported newly diagnosed conditions in our study were tachycardia, hypertension, and POTS. There is growing evidence of POTS diagnoses following COVID-19; a cross-sectional study of COVID-19 survivors reported a 19% prevalence of POTS diagnosis in 802 survivors.^22^ The implications of these results include the substantial challenges for individual with PASC and increased utilization of already strained health resources, systems, and healthcare workers.^26^ This also highlights the urgent need for coordinated and integrated transdisciplinary approaches towards management, and both population level and individual level interventions to promote recovery.

Cardiovascular assessment and development of diagnostic algorithms for patients with PASC would be beneficial, particularly for patients not previously hospitalized. Mobile heart rhythm tracking devices and Holter ECG monitoring for detecting arrhythmias and abnormal pulse reactions, and assessment of heart rate are beneficial non-invasive outpatient diagnostic strategies to further characterize symptomatic tachycardia and other cardiac symptoms associated with PASC.^29^ There is a need for specific recommendations and policies to help with diagnosis and management of PASC, particularly considering the anticipated fiscal impact of PASC on population productivity. As mechanisms for enduring cardiac injury post-acute COVID-19 are poorly understood, additional investment in under-resourced communities most hit by COVID-19 infections and PASC is critical.^12^ Investment in cardiac-related PASC research is equally important to inform potential therapeutic strategies for PASC. In addition, there are urgent needs for coordinated efforts directed at resource allocation and optimization of primary care for persons with cardiac PASC and for prevention of resulting CV events and complications.^23^ Although several tertiary medical centers have introduced post-acute COVID clinics, referrals are not readily accessible, further underscoring the importance of investing in primary care for equitable PASC care.^12^ This includes the need for population-level strategies to address the rising burden of CVD attributable to COVID-19.

The findings, particularly the prevalence of symptoms and associated CV morbidity, suggests the critical need for access to PASC care, including better understanding of the management of cardiac PASC. Finally, this study addresses research priorities for understanding the cardiac-related PASC symptom burden, assessing the impact of these lingering symptoms in persons with pre-existing CV risk factors, CVD, and the future risk of complications.

## Limitations

Lack of temporality is a major limitation in our observational study; hence, causal inference cannot be ascertained. Another limitation is that our data were based on self-report with non-ascertainment of reported CVD diagnoses or reported symptoms through EMR chart review. Self-report is an acceptable method of data collection in epidemiological studies, and may be used as a proxy in the absence of EMR data.^30^ Also, most of our participants were never hospitalized for COVID-19 and have yet to encounter health systems concerning their conditions. Considering the current uncertainty regarding diagnostic criteria for PASC and cardiac-related PASC, we intend to further assess diagnosed cardiac conditions using EMR ascertainment as the next step in this study. Additionally, since the data were self-reported, we recognize this may have been subject to subjective symptoms report and recall bias. Another limitation to note is the possibility of residual confounding. We accounted for measured confounding; however, residual confounding due to unmeasured factors is another limitation of our study.

Nevertheless, our study has several strengths. First, the majority of our sample includes non-hospitalized patients, community-dwelling persons with PCR confirmed COVID-19 diagnosis tested at the Johns Hopkins system in our cohort, unlike many other studies based on hospitalized patients. Also, we recruited participants interested in joining COVID-19 studies from a COVID-19 registry with a response rate of 70%. Additionally, we provided information on cardiac-specific symptoms by evaluating all cardiac PASC symptoms as a group, rather than as individual symptom outcomes.

## Conclusion

In conclusion, among the sample of community-dwelling adults diagnosed with COVID, we estimated the burden of possible cardiac-related PASC symptoms to be 43%, and the diagnosed cardiac conditions including hypertension was around 27%. Our findings suggest that underlying CVD, pre-existing conditions, older age, COVID-19 hospitalization, and ≥18 months since testing positive may be risk factors for persistent cardiac-related PASC symptoms. Hence, COVID-19 may play an important role in worsening the prognosis of existing CVD and pre-existing conditions and in increasing the risk of complications.

## Data Availability

All data produced in the present study are available upon reasonable request to the authors

## Acknowledgment

We would like to thank the Cardiac PASC study participants. We also thank Cassie Lewis-Land, Scott Carey, Mike Sherman, and the Institute of Clinical and Translational Research (ICTR) Recruitment Innovation Team for technical support during data collection.

## Keywords and Non-standard Abbreviations and Acronyms

PASC: Post-Acute Sequelae of SARS-CoV-2 Infection
COVID-19: Coronavirus Disease 2019
CVD: cardiovascular disease
SARS-CoV-2: severe acute respiratory syndrome coronavirus 2

