## Supplemental Table 1 for "Cardiac Post-acute Sequelae symptoms of SARS-CoV-2 in Community-Dwelling Adults: Cross-sectional Study"

**Supplemental Materials**

Supplemental Table 1. Newly diagnosed conditions after acute COVID-19 infection stratified by time since infection

| Newly diagnosed Conditions, n(%) | Total N=119 | Time since infection | | *P*-value |
|---|---|---|---|---|
| | | <18 Months (N=81) | ≥18 Months (N=38) | |
| Hypertension | 24 (20.2) | 18 (22.2) | 6 (15.8) | 0.415 |
| Tachycardia | 29 (24.4) | 14 (17.3) | 15 (39.5) | **0.009** |
| Myocarditis | 5 (4.2) | 1 (1.2) | 4 (10.5) | **0.019** |
| Cardiomyopathy | -- | -- | --- | -- |
| Myocardial injury | 1 (0.8) | -- | 1 (2.6) | 0.143 |
| Atrial fibrillation or flutter | 8 (6.7) | 4 (4.9) | 4 (10.5) | 0.256 |
| Pericarditis | 1 (0.8) | 1 (1.2) | -- | 0.492 |
| Myocardial infarction | 1 (0.8) | 1 (1.2) | -- | 0.492 |
| Heart block | 3 (2.5) | 2 (2.5) | 1 (2.6) | 0.958 |
| Heart failure | 3 (2.5) | 2 (2.5) | 1 (2.6) | 0.958 |
| Pulmonary embolism | 1 (0.8) | -- | 1 (2.6) | 0.143 |
| Deep Vein Thrombosis | 2 (1.7) | 1 (2.6) | 1 (2.6) | 0.580 |
| Stroke | 1 (0.8) | 1 (1.2) | -- | 0.492 |
| Acute kidney disease | 2 (1.7) | -- | 2 (5.3) | 0.037 |
| Postural orthostatic tachycardia syndrome (POTS) | 16 (13.5) | 8 (9.9) | 8 (21.1) | 0.096 |

Bold: Statistically significant at <0.05. P-values estimated using chi-square test statistics for differences in proportions.
