## Supplementary figures and images for "Cardiac Post-acute Sequelae symptoms of SARS-CoV-2 in Community-Dwelling Adults: Cross-sectional Study"

### Supplemental Figure 1

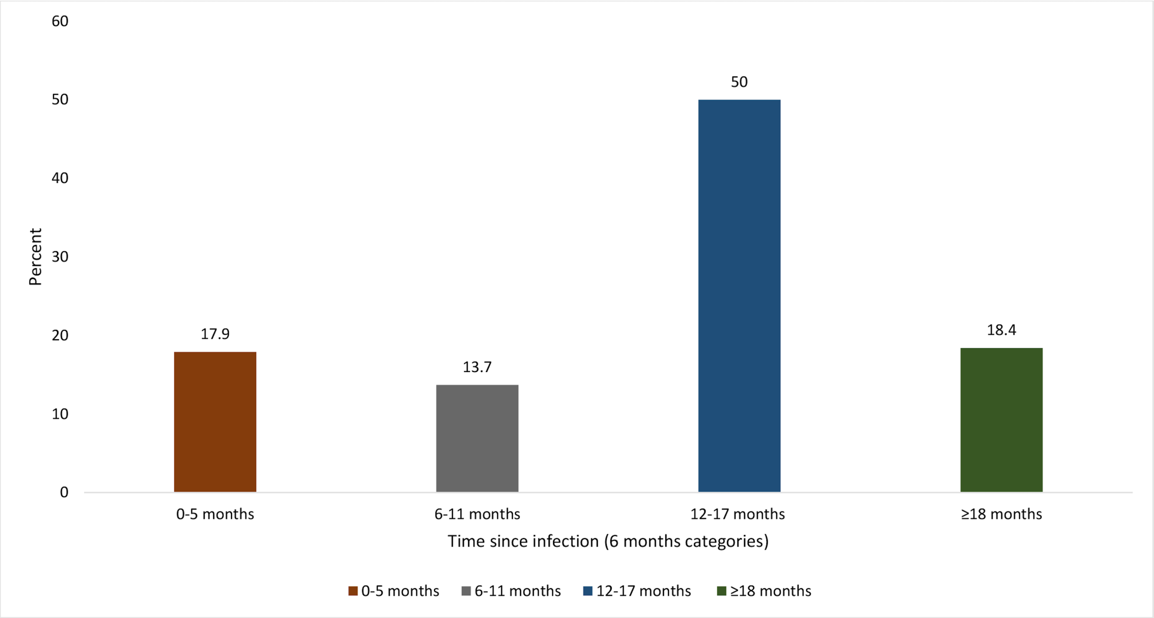
